# Life-long effects of malnutrition using semi-quantitative EEG analysis

**DOI:** 10.1101/2022.01.18.22269447

**Authors:** Fuleah A. Razzaq, Ana Calzada Reyes, Qin Tang, Yanbo Guo, Yujie Liu, Lidice Galan-Garcia, Anne Gallagher, Trinidad Virues-Alba, Carlos Suarez-Murias, Arielle Rabinowitz A., Ileana Miranda, Vivian Bernardo Lagomasino, Janina R Galler, Maria L. Bringas-Vega, Pedro A. Valdes-Sosa

**Affiliations:** The Clinical Hospital of Chengdu Brain Science Institute, MOE Key Lab for Neuroinformatics, University of Electronic Science and Technology of China, Chengdu, China; Cuban Neuroscience Center, La Habana, Cuba; LION Lab, Sainte-Justine University Hospital Research Centre, University of Montreal, Montreal, QC, Canada; Department of Neurology and Neurosurgery, McGill University, Montreal, QC, Canada; National Center for Animal and Plant Health, CENSA, Mayabeque, Cuba; Facultad de Psicologia, Universidad La Habana, La Habana, Cuba; Division of Pediatric Gastroenterology and Nutrition, Mass General Hospital for Children, Boston, MA, United States of America

**Keywords:** Malnutrition, EEG, latent variable, item-response theory, semi-quantitative analysis, GTE scale

## Abstract

1

The non-linear spatiotemporal features in the continuing EEG recordings could be helpful to infer the physio and pathological significance of early insults on the brain, such as early malnutrition and their long-term effects. A unique opportunity is opened with the Barbados Nutrition Study (BNS) dedicated to studying Protein-Energy Malnutrition (PEM) with two groups, children suffering an early PEM episode and their controls. We evaluated the resting-state EEG (N=108, PEM=46) in 1978, and we repeated the EEG (N=97, PEM=46) in 2018. We did a qualitative analysis of the EEG using a semi-quantitative scale (Grand Total EEG (GTE)) and an item response theory (IRT) approach to estimate a latent variable that is able to explain the subjacent neurophysiological status (NPS). Finally, we applied a mixed-effects model with a sensitivity index for ignorability to test differences between the controls and PEM groups while accounting for the missing data mechanisms (*nlme* (Pinheiro J. 2020) and the *ISNI* package in R(Xie et al., 2018). The fixed effects were group, age, gender, and socioeconomic status; the random effect was the variability inherent to each participant and evaluator.

**Results:** The simple visual inspection of the 1978 EEG recordings detected 39 participants with abnormalities (28 PEM and 11 Controls; p<0.05); in 2018, a total of 63 participants showed abnormalities in the EEG recordings (35 PEM and 28 Controls; p<0.01)).

The polytomous IRT analysis revealed that all items had been loaded well onto the latent factor, and the highest value of the Focal abnormality reached 0.97. The fixed effect of the groups (PEM vs. Control) was highly significant, with a **p-value of 0** and the **c index of 5.27**. Age was also significant with a **p-value of 0.0093** and the **c index of 14.793**, whereas Gender and SES were not significant. The contrasts at the two different time points (childhood (1978) mean age= 8.45, adulthood (2018) mean age=48.30) also showed highly significant differences between groups with a **p-value of 0**.

**Conclusions:** EEG abnormalities were seen in both PEM and control groups during the school years and later in middle adulthood, with a higher proportion of abnormalities in the previously malnourished BNS participants at both ages. The statistical significance of these differences was confirmed through a latent variable approach and a linear mixed-effect model, which discriminated successfully against the long-term effects of early malnutrition on the brain up to 50 years after the onset of malnutrition in the first year of life.

## 1. Introduction

The electroencephalogram (EEG) is the most used neuroimaging technique to identify the effects of early insults on the brain, including protein-energy malnutrition (PEM). For a recent review, see Galler et al. 2021. The EEG analysis uses either qualitative or quantitative methods, with each approach focusing on different properties and features of the EEG recordings. The qualitative analysis evaluates the interictal EEG, which is related to the recordings that do not contain seizures or ictal manifestations, and it is the most frequent recording type in clinical practice. The interictal EEG recordings are characterized by non-stationary and non-linear features of the electrophysiological activity, such as the graph-elements (sharp-waves, spikes, spike and wave, polyspikes, polyspikes, and waves), which are transient with a waveform determined and the slow activity (focal or generalized) and paroxysmal activity. All of them could be easily detected by simple visual inspection of experts neurophysiologists following the glossary for clinical electroencephalographers (Kane, F 2017). On the other hand, the quantitative analysis is based on the lineal and stationary properties of the EEG across the EEG spectrum and autoregressive time series tools, which do not include the graph elements mentioned before.

Our group demonstrated that the results from both methods are complementary based on concurrent validation in a recent cross-sectional study where we found differences between EEG childhood in participants from the Barbados Nutrition Study (BNS) who suffered an early episode of PEM compared to healthy controls. (Taboada et al., 2018).

Longitudinal studies constitute the best methodological approach to study how malnutrition can influence brain function. Accordingly, we report here the qualitative and semi-quantitative analysis of EEGs recordings done in 1978 and 40 years later, 2018. We used visual inspection of EEG recordings by two experts to identify abnormalities and score the individual items of the one semi-quantitative scale, Grand Total EEG (GTE), which has been employed to study EEG abnormalities in Dementia and Parkinson disorders with good sensitivity and specificity and the discrimination between other pathologies, frontotemporal lobar degeneration, Dementia Lewis bodies and/or stages of disease severity. (Lee, Brekelmans, & Roks, 2015; Micanovic & Pal, 2014;Pijnenburg et al., 2008; Roks, Korf, Van Der Flier, Scheltens, & Stam, 2008). Contrary to previous studies, we utilized an item response theory approach instead of the total score to analyze and interpret the results. First, we examined the values of the raw items to obtain their reliability to explain the underlying relations with the experts’ assessment (observations). Later a polytomous IRT was implemented to get the latent factor scores for the GTE scale using the *mirt* package in R (Chalmers 2012). Moreover, a mixed-effects model was applied with a sensitivity index for ignorability to test differences between control and PEM groups while accounting for missing data mechanisms using *nlme* (Pinheiro J. 2020) and *ISNI* package in R(Xie et al., 2018). The fixed effects were group, age, gender, and socioeconomic status, and the random effect was the variability inherent to each participant and the two evaluators.

The present study aimed to determine whether visual and EEG semi-quantitative analysis using a latent variable can shed light on the long-term effects of childhood malnutrition.

## 2. Material and Methods

### 2.1 Study Site

The current study was conducted in Barbados, a Caribbean country whose population is 280,000. The demographic makeup is 92% African/Caribbean origin, 4% Caucasian, and 4% individuals of Asian, Lebanese, and Syrian descent. In 1970, the infant mortality rate was 46 per 1,000 live births. Today it stands at 7.8, and Barbados is ranked as 52 on the Human Development Index (United Nations Development Program [UNDP], 2016). Moderate-severe cases of infant malnutrition were of significant concern when this study was undertaken in the 1970s. Nowadays, infant malnutrition is virtually eliminated from the island due to its improved economy and the impact of island-wide nutrition-related education.

The Barbados Nutrition Study (BNS) has been following the original first generation of this sample for more than 50 years. More information about the original study can be found in (Galler et al. 1983a,b, Ramsey 1979) and more recent results in (Taboada-Crispi et al., 2018; Galler et al., 2013; Galler et al., 2012; Hock et al., 2017; Hock et al., 2020). Study participants (PEM) were born between 1967-1972 and had a history of childhood malnutrition (weight lost (below 75% of expected weight for age, in the absence of edema) limited to the first year of life. The inclusion criteria were significant; their birth weight was at least 5 lbs.; no evidence of prenatal or prenatal complications, Apgar scores >8, and no history of convulsions, head injury, or loss of consciousness. The healthy controls (CON) were classmates of the PEM group with no malnutrition or other severe medical conditions and were matched to the PEM group by age, gender, and handedness.

### 2.2 Samples

The first EEG dataset of the original BNS sample was recorded between 1977-1978 (n=258) when the subjects were between 5-12 years old, and 108 EEG recordings were valid and used for subsequent analysis. EEG was re-recorded 40 years later in 2018 (n=97) when the participants were between 45-51 years with 94 valid EEG recordings (mean age was 48.2 years (SD 2.14). Note that 55 subjects dropped out of the study and do not have EEG in the second wave (2018). Whereas 53 subjects had EEGs at both waves (1978, 2018), and the remaining 41 participants had the EEG missing at the first time (1978).

It’s important to note that the original size sample decreased from the original study in the ‘70s to the final EEG recordings in 2018. This attrition of the original sample was due to different causes: participants living overseas (17), others did not respond to the call (16) or refuse (14) to participate; other six were deceased, one in prison, and other severely ill. To confirm if the missing cases of EEG affected the results presented here, we implemented a sensitivity analysis for the ignorability assumption, detailed in the statistical analysis section.

### 2.3 Ethics

The Barbados Ministry of Health granted general approval for this study, the Massachusetts General Hospital IRB (2015P000329/PHS/MGB), and the Cuban Neuroscience Center (2017/02/17/CNEURO). All the participants were volunteers and signed informed consent before the study. They were compensated for their time and travel to and from the Barbados Nutrition Study (BNS) Center.

### 2.4 EEG procedure

The original 1978 EEG protocol is described in detail in Taboada et al. (2018).

The 2018 follow-up EEG recordings were performed using the same procedures as those in the earlier study with a 21-channel digital EEG hardware and software package (MEDICID 5, Neuronic, Havana, Cuba).

Surface electrodes were placed at 19 sites (Fp1, Fp2, Fz, F3, F4, F7, F8, Cz, C3, C4, T3, T4, T5, T6, Pz, P3, P4, O1, and O2) according to the international 10-20 system, and they were referenced to linked earlobes with a ground electrode attached to the forehead. The impedance of all electrodes did not exceed five kΩ throughout the recordings. EEG signals were amplified 10,600-fold, bandpass filtered from 0.5 to 30 Hz, and sampled by a 12-bit analog-to-digital converter at 200 Hz.

Resting-state EEG was recorded for 8-10 minutes with the eyes closed, followed by two minutes of closed and opened eyes sequences, three minutes of hyperventilation, and two minutes of recovery. The individual vigilance level was checked during EEG recordings to monitor the slowing of the EEG background activity or the appearance of sleep patterns.

#### 2.4.1 Visual EEG assessments

The EEG recordings were evaluated independently by two clinical neurophysiologists from the Cuban Neuroscience Center (ACR, TVA) who were blind to the participant’s malnutrition history, using the standard criteria from the International Federation of Clinical Neurophysiology (Babiloni et al., 2020).

In general, the EEG recording was considered normal if an adequate organization of background activity (as per the subject’s age), well-defined spatial differentiation, rhythmic alpha-band activity, and the absence of paroxysmal activity were present. EEG abnormalities were defined as spikes, sharp waves, periodic discharges, triphasic waves, and intermittent slowing in the EEG activity.

The slow-wave activity was considered as the presence of persistent nonrhythmic theta-and delta-band slow waves. In contrast, paroxysmal activity was characterized by a sudden onset, rapid attainment of a maximum, and abrupt termination, and distinguished from background activity such as a spike, a sharp wave, and a spike and wave. A spike is by convention defined as a waveform with a duration between 20 and 70 msec. Paroxysmal activity that lasts 70-200 milliseconds is referred to as a sharp wave and spike and a wave complex if a delta-band wave follows one spike.

For a detailed description and referral of the scalp EEG waveforms and physiological and pathophysiological EEG graph-elements in adults and children, see the IFCN and EFNS Guidelines (Beniczky et al., 2017; Waldemar et al., 2007).

### 2.5 GTE Scale

To carry out a more sensitive analysis of the EEG visual inspection data, we devised a Likert-type scale based on an ordinal grading scale of perceived abnormality (modified from Jonkman, 1989 and de Weerd et al., 1990). This semi-quantitative scale summarized the EEG findings of the EEGs previously found by visual inspection, where 0 is the absence of abnormality and 1, 2, or 3 represent different severity levels. A total score was not used. Instead, we used item analysis to estimate the latent variable underlying scale.

The original GTE scale was composed of 6 items1:

1. Frequency of rhythmic background activity
2. Slow activity
3. Reactivity of rhythmic background activity
4. Paroxysmal activity*
5. Focal abnormality
6. Sharp wave activity

But we only selected the last five items for the analysis, eliminating the third one because the recordings in 1978 only included the closed eyes condition, which prevented us from analyzing the reactivity.

**For the 2018 analysis, we included additional modifications:**

a. Item 2 (Slow theta), response options 4 and 5 were excluded from the original GTE scale.
b. Item 4 (Paroxysmal activity) option 0 was assigned to EEG without paroxysmal activity, 1 when EEG had paroxysmal slow activity, 2 when EEG had spikes, and three spikes and waves.
c. Item 5 (Sharp wave activity) we excluded triphasic wave and PLEDS for similar reasons stated above.
d. Unlike the previous study in 1978, we included item 3, “Reactivity of rhythmic background activity,” because we measured the EEG during different functional states (closed-eyes, open-eyes, and hyperventilation), which allowed the assessment of the reactivity. Note that in 1978 the recordings only included closed-eyes.
e. *Note that the term “paroxysmal activity” is the original but has not found general acceptance. For the discussion of the results, we will employ the term interictal-epileptiform discharge IED, even when this term also has been attacked on the ground that such discharges may occur in the absence of clinical seizures manifestations or in individuals who have never had seizures.

### 2.6 Statistical analysis

1. Chi-square contingency tables were used to analyze the statistical significance of the frequency of abnormalities detected by visual inspection of the EEG recordings of both groups in 1978 and 2018.
2. We performed an Item Response Theory (IRT) analysis on the GTE scale. IRT is used when the indicator variables are categorical and not continuous. This method identifies the most informative items and obtains their optimal linear combination using non-linear factor analysis to produce a final score (f1). This score represents a “latent variable,” which we refer to as “neurophysiological status” (NPS) for the EEG (see Taboada et al., 2018). It is essential to point out that the selection criteria based on high factor scores are geared towards picking items that show a clear separation of probabilities between the different levels of the scale and *not* between the two groups (control vs. PEM) since responses for all participants were included in this analysis. The latent variable was estimated using the original raw items scores (observations): five common items for both the 1978 and 2018 EEG dataset, using the sample of N=202 (108 samples from 1978 and 94 from 2018). The GTE scale range from 0-5. We have implemented a polytomous IRT (Beaujean, 2014; P. Chalmers, 2015) using the R package MIRT (R. P. Chalmers, 2012). We constructed a latent factor using a generalized partial credit model (R. P. Chalmers, 2015;
3. Pollitt & Hutchinson, 1987). For the initial model, we included five common items. We assessed the model based on item loadings and trace plots. We further optimized the initial model and compared it based on model fit indices like (AIC, BIC).
4. The second step was to integrate the latent variable in the linear mixed effect (LME) model to test the differences between groups. We select the function *lme* from the *nlme* R package (Pinheiro J. 2020). We analyzed age and group for fixed effects while adjusting for gender and childhood socioeconomic status (SES) whereas, subjects and evaluators were used as random effects under different models, and the model with better fit indices was selected.
5. We performed a sensitivity analysis for the ignorability assumption. The ignorability assumption states that all the confounding factors should be adjusted to infer from the regression coefficient as average effects. Thus, making the group/treatment assignment ignorable, just like a completely randomized experiment. However, the missing data mechanism Missing Not At Random (MNAR) can violate this assumption. To assess any effects of missing data mechanisms on the LME model estimates, we applied an index of local sensitivity to non-ignorability (ISNI) using the R package ISNI (Ma, Troxel, & Heitjan, 2005; Xie et al., 2018). ISNI provides a standardized sensitivity transformation statistic “c” where a large c means that LME estimates are robust, and only extreme violation to ignorability assumption can change the initial estimates. Thus, non-ignorability is of little concern. A rule of thumb by (Troxel et al. 2004) is c >= 1 shows robust estimates.

## 3 Results

### 3.1 Qualitative analysis of the EEG

#### EEG abnormalities reported by visual inspection

Visual inspection of the EEG recordings demonstrated a preponderance of abnormalities in the PEM group compared to the controls in both 1978 and 2018 datasets, with more abnormal findings reported in 2018 than in 1978.

**1978:** The total number of EEG recordings with abnormalities was 39/108, with 28 (60.9%) belonging to the PEM group and 11 (17.7%) to the control group. The chi-square statistic is 6.5377. (p-value is 0.010561).

**2018:** The total of EEG recordings with abnormalities was 63/98, with 35 (77.7%) in the PEM group and 28 (52.7%) in the Control group (p<0.001). The chi-square statistic with Yates correction is 5.7877 (p-value 0.016138).

### 3.2 IRT analysis

The initial IRT model with five items showed that the Background Frequency is not associated with the latent factor with an f1 score of 0.134. Furthermore, the trace plots or Item Characteristic Curve (ICC) for Sharp waves showed that “Sporadic sharp waves” are not informative and are rarely used. We observed similar trends in the trace plot for paroxysmal activity, which showed a binary trend with only two responses being used by evaluators “0=None” and “1=Paroxysmal activity/spikes”. The model fit indices showed AIC 3424.02 and BIC 3508.21. We optimized the model based on these findings by excluding background frequency and collapsing the response categories for Sharp waves and Paroxysmal activity. This resulted in a model with better fit indices with AIC 2494.16, BIC 2554.29. The results of the optimized model are in Table 1, which shows the f1 scores for each item. All items are loading well onto the latent factor, and Focal abnormality has the highest value of 0.97. Figure 1 shows the trace plots for each item, showing a curve for every response category under each item and representing it against the latent factor theta. The trace plots for the optimized model show good trends.

**Table 1.** The IRT results for the GTE scale. Note that “Focal abnormality” and “Paroxysmal activity” were the most significant items to discriminate between groups (F1>0.7)

| Items | F1 |
| --- | --- |
| Diffuse slow activity | 0.652 |
| **Paroxysmal activity** | **0.726** |
| **Focal abnormality** | **0.976** |
| Sharp waves | 0.563 |

**Figure 1.**
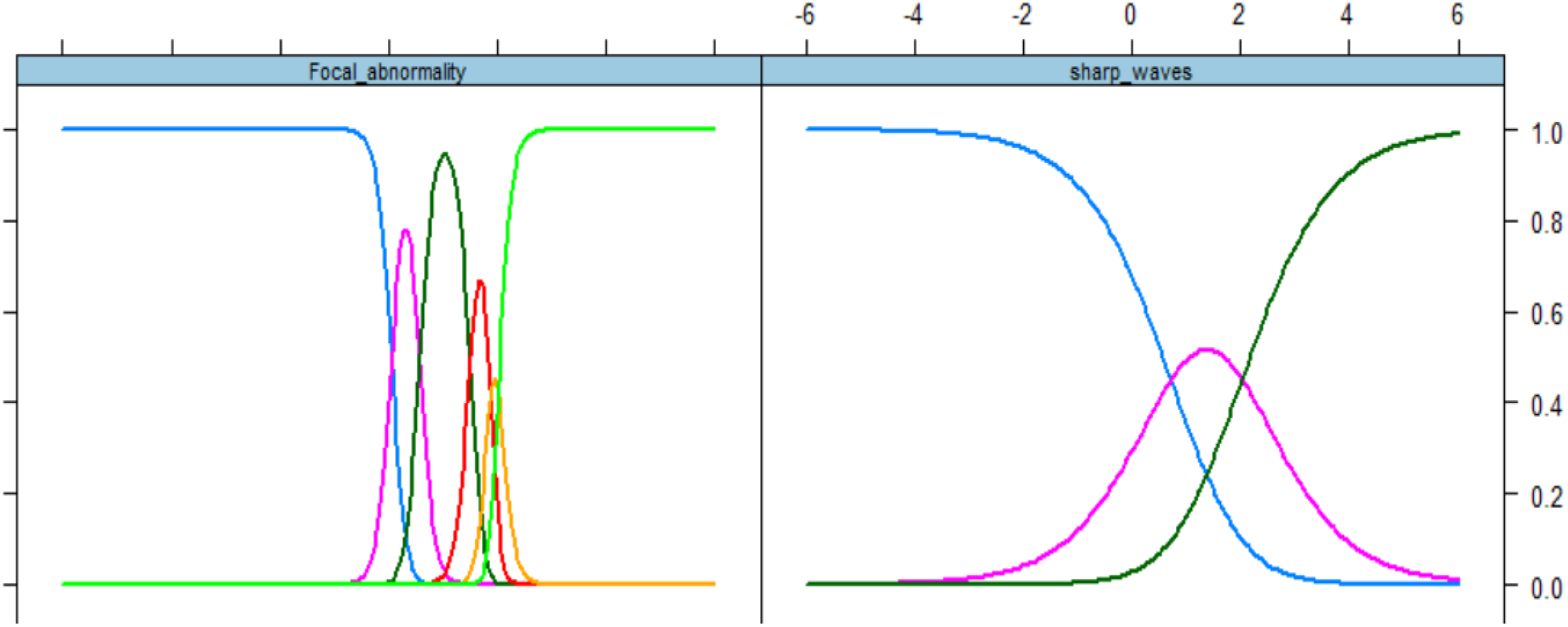
Trace/ICC plots. The x-axis is the value of the latent variable (*θ*), and the y-axis is the probability *P*(*θ*) that shows the chance of occurrence for each response category across different levels of *θ*. Top Left: Focal Abnormality, the item with the best discrimination (0.97) Top Right: Sharp wave the item with worst discrimination (0.53).

### 3.3 Mixed-effects model to compare the two groups

The model employed to test the differences between groups is

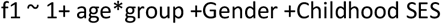

We tested for random effects of subjects and evaluators. The mixed-effect model with the smallest AIC and BIC values only had the subjects as a random effect. So, we used that model for inference. The fixed effect of the group was highly significant, with a **p-value of 0** and the **c index of 5.27**.

Age was also significant with a **p-value of 0.0093** and the **c index of 14.793** whereas, Gender and SES were not significant. The contrasts at different time points (childhood (1978) mean age= 8.45, adulthood (2018) mean age 48.30) also showed highly significant differences between groups with a **p-value of 0**.

In our results, we obtained an index **c >= 1**, indicating that the analysis was robust and the missing EEG data was not significant.

## 4 Discussion

### Comparison between qualitative and quantitative EEG analysis

With the development of qEEG, the qualitative and/or semi-quantitative analysis of the EEG is less represented in the scientific literature. Even if the quantitative EEG has been employed repeatedly and successfully to demonstrate differences between groups, we recognize that the other non-linear and non-stationary features of the EEG, which provide rich information about the underlying electrophysiological changes in these differences, are not considered. Note that the conventional EEG analysis is based solely on the visual examination of the continuous tracings, which may be highly subjective and therefore a disadvantage. Nevertheless, neurophysiologists can provide unique and valuable information about EEG resting state.

The standard input for the quantitative EEG analysis is the artifacts-free segments of EEG based on spectral analysis. These are selected by visual inspection during the revision of the EEG, and they are supposed to be quasi-stationary. For that reason, other non-linear features of the EEG are missing in the analysis. Although the interpretation of the EEG by visual inspection depends mainly on the experience, knowledge, and subjective judgment of the neurophysiologist, it is essential to note that in our studies, we did not find differences between the results obtained by the area under the curve (automated operator) and the results reported by the clinical neurophysiologists, as published with the EEG of 1978 (Taboada et al.,2018).

### Prevalence of findings to compare PEM vs. Control using GTE scale

The GTE scale results were more informative when an item analysis was employed rather than a global score. The item-response theory approach used in this report was a robust and reliable method to identify the differences between nutrition groups in childhood and middle adulthood. The persistence of “focal abnormalities” appeared to be the most discriminant item supporting the differences between groups at both time points.

Note that the focal abnormality item includes both focal slow activity and/or focal paroxysmal activity.

a. The focal slow abnormalities are related to the increment of delta and theta rhythms in the recordings, which is the contrary effect expected for a normal EEG maturational process. (Segalowitz et al.,2010;Uhlhaas et al., 2010). The persistence of slow focal abnormalities could represent the impact of early childhood malnutrition on adult brain function.
b. The focal paroxysmal activity has a prevalence in normal children from 0.8 to 18.6% and 0.3 to 12.3% in normal adults, reported by (Shelley 2008) in a meta-analysis of 22 papers between 1950 and 2005, which summarized more than 50,000 subjects.

The prevalence rates of IED reported in patient populations are generally higher than those of healthy subjects. (Sam and So, 2001) described a range between 2 −12.3% in the neurological patient (inpatients and/or outpatient) found that around three-fourths of non-seizure patients with IED had acute or progressive brain disorders. (Shelley, 2008) in the same metanalysis, reported a high incidence of paroxysmic activity in neuropsychiatric disorders such as schizophrenia (30-60%), 60% in a panic attack and OCD, mood disorders 20-40%, personality disorders 10-53%.

In our study, the prevalence of focal abnormalities is very high in both EEG evaluations.

For example, in 1978, during childhood, the PEM group showed 73.9% of participants with focal abnormalities, of which 21.4% were paroxysmal activity. On the other hand, in the control group, the incidence of paroxysmal activity was 14.3%, lower than the PEM group and lower than the range reported by Shelley (2008) for healthy children (08.-18.6 %).

But in 2018, the PEM group exhibited 18.6% of paroxysmal activity during adulthood and the control group 14.4%, both groups higher than the prevalence range of 0.3 to 12.3%, reported by (Shelley 2008) for normal adults.

These electrophysiological abnormalities may reflect central nervous system dysfunction at the cortical and neuronal levels or may be a consequence of neurochemical modifications associated with the history of early malnutrition.

The diffuse slow activity was another critical item that showed a significant difference between both groups. This type of EEG abnormality is related to global cerebral dysfunction. This finding could be signaling the irreversible neurological consequences of early childhood malnutrition (Galler, 1984; Galler et al., 1996).

Differences between groups.

The *lme* analysis demonstrated that the fixed effect of the group was highly significant, with a **p-value of 0** and the **c index of 5.27**. This global result was tested separately using contrasts for childhood and adulthood. Age was also significant with a **p-value of 0.0093** and the **c index of 14.793** whereas, Gender and SES were not significant.

Age-related changes in the EEG are consistent with known neurobiological and neuroanatomical changes that occur during typical aging. This strong influence of age on brain electrical development has been published elsewhere. (Niedermeyer, Handbook Clinical Neurophysiology).

On the other hand, the relation of gender and socioeconomic status on the EEG has not been well demonstrated, with contradictory results mainly related to the methodology employed. In our previous analysis of the BNS childhood EEG dataset, we didn’t find any relation with gender (Taboada 2018, Bringas 2019).

Socioeconomic status (SES) is a multifactorial variable composed of environmental, social, and economic factors influencing the behavior and development of the subjects, which has been scarcely employed in EEG longitudinal studies. Note that in our case, in the childhood EEG study, the instrument evaluated parent’s SES and, in adulthood, was the participant SES.

In this study, one limitation could be the limitations of the original sample of participants with EEG in 1978. However, the sensitivity procedure we employed to test the influence of missing data in the analysis discarded this effect on the results.

## 5 Conclusions

Our findings demonstrate that visual inspection and semi-quantitative EEG analysis are efficient and reliable tools to establish differences between previously malnourished and control groups. This approach can provide feasible and cost-effective tools to the general neurologists in underserved clinical settings where semi-quantitative scales are available.

## Data Availability

All data produced in the present study are available upon reasonable request to the authors

## 6 Contribution of the authors

Conceptualization: MLB, PAVS

Data acquisition: ACR, CSM

Data curation, scoring GTE: ACR, TVA, YB, QT, VBL

Methodology: PAVS, IM, LGG, MLB, FAR

Statistical Analysis: FAR

Funding acquisition: MLB, PAVS, JRG

Project administration: MLB, JRG

Supervision: AG, JRG

Writing original draft: MLB, ACR, PAVS

Writing-review-editing: FAR, XL, YL, AR, JRG, PAVS

## 7 Acknowledgments

We would also like to express our gratitude to the Barbados Nutrition Study staff, with special thanks to Alicia Innis and Dr. Cyralene Bryce. We would also like to thank the dedicated study participants and their families who have participated in this study over their lifetimes. This research was conducted in cooperation with the Ministry of Health of Barbados and was supported by grants from Nestle Foundation (to MLB and PAVS, Validation of a long-life neural fingerprint of early malnutrition, 2017), National Institutes of Health (HD060986 to JRG), the NSFC Project No. 61871105) and CNS Program of UESTC (No. Y0301902610100201).

## Notes

### Competing Interest Statement

The authors have declared no competing interest.

## References

Babiloni, C., Barry, R. J., Başar, E., Blinowska, K. J., Cichocki, A., Drinkenburg, W. H. I. M., … Nunez, P. (2020). International Federation of Clinical Neurophysiology (IFCN)–EEG research workgroup: Recommendations on frequency and topographic analysis of resting state EEG rhythms. Part 1: Applications in clinical research studies. Clinical Neurophysiology, 131(1), 285–307.

Beaujean, A. A. (2014). Latent variable modeling using R: A step-by-step guide. In Latent Variable Modeling Using R: A Step-by-Step Guide. https://doi.org/10.4324/9781315869780

Beniczky, S., Aurlien, H., Brøgger, J. C., Hirsch, L. J., Schomer, D. L., Trinka, E., … Herman, S. T. (2017). Standardized computer-based organized reporting of EEG: SCORE – Second version. Clinical Neurophysiology. https://doi.org/10.1016/j.clinph.2017.07.418

Chalmers, P. (2015). Item response theory Unidimensional IRT Multidimensional IRT Diagnostics Estimation Package Specifics Multiple Group IRT, DIF, and DTF Multidimensional Item Response Theory Workshop in R.

Chalmers, R. P. (2012). mirt : A Multidimensional Item Response Theory Package for the R Environment. Journal of Statistical Software, 48(6), 1–29. https://doi.org/10.18637/jss.v048.i06

Chalmers, R. P. (2015). Extended Mixed-Effects Item Response Models With the MH-RM Algorithm. Journal of Educational Measurement, 52(2), 200–222. https://doi.org/10.1111/jedm.12072

De Weerd, A., Perquin, W., and Jonkman, E. (1990). Role of the EEG in the prediction of dementia in Parkinson’s disease. Dementia 1, 115–118. doi: 10.1159/000107129

Galler, J. R., Bringas-Vega, M. L., Tang, Q., Rabinowitz, A. G., Musa, K. I., Chai, W. J., Omar, H., Abdul Rahman, M. R., Abd Hamid, A. I., Abdullah, J. M., & Valdés-Sosa, P. A. (2021). Neurodevelopmental effects of childhood malnutrition: A neuroimaging perspective. NeuroImage, 231, 117828.https://doi.org/10.1016/j.neuroimage.2021.117828

Galler, J.R., Bryce, C.P., Zichlin, M.L., Waber, D.P., Exner, N., Fitzmaurice, G.M. and Costa, P.T., 2013. Malnutrition in the first year of life and personality at age 40. Journal of Child Psychology and Psychiatry, 54(8), pp.911–919.

Galler, J.R., Bryce, C.P., Zichlin, M.L., Fitzmaurice, G., Eaglesfield, G.D. and Waber, D.P., 2012. Infant malnutrition is associated with persisting attention deficits in middle adulthood. The Journal of nutrition, 142(4), pp.788–794.

Galler, J. R., Ramsey, F., Solimano, G., Lowell, W. E., & Mason, E. (1983)a. The Influence of Early Malnutrition on Subsequent Behavioral Development: I. Degree of Impairment in Intellectual Performance. Journal of the American Academy of Child Psychiatry, 22(1), 8–15. https://doi.org/10.1097/00004583-198301000-00002

Galler, J. R., Ramsey, F. C., Solimano, G., & Lowell, W. E. (1983)b. The Influence of Early Malnutrition on Subsequent Behavioral Development II. Clasroom Behavior. Journal of the American Academy of Child Psychiatry, 22(1), 16–22.

Hock, R.S., Bryce, C.P., Waber, D.P., McCuskee, S., Fitzmaurice, G.M., Henderson, D.C. and Galler, J.R., 2017. Relationship between infant malnutrition and childhood maltreatment in a Barbados lifespan cohort. Vulnerable children and youth studies, 12(4), pp.304–316.

Hock, R.S., Rabinowitz, A.G., Bryce, C.P., Fitzmaurice, G.M., Costa Jr, P.T. and Galler, J.R., 2020. Intergenerational effects of childhood maltreatment and malnutrition on personality maladaptively in a Barbadian longitudinal cohort. Psychiatry Research, p.113016.

Jonkman, E. A. (1989). Simple EEG scoring method for senile dementia of the Alzheimer type. Electroenceph. Clin. Neurophysiol. 72:44.

Kane, N., Acharya, J., Benickzy, S., Caboclo, L., Finnigan, S., Kaplan, P.W., Shibasaki, H., Pressler, R., van Putten, M.J.A.M., 2017. A revised glossary of terms most commonly used by clinical electroencephalographers and updated proposal for the report format of the EEG findings. Revision 2017. Clin. Neurophysiol. Pract. 2, 170–185. https://doi.org/10.1016/j.cnp.2017.07.002

Lee, H., Brekelmans, G. J. F., & Roks, G. (2015). The EEG as a diagnostic tool in distinguishing between dementia with Lewy bodies and Alzheimer’s disease. Clinical Neurophysiology, 126(9), 1735–1739. https://doi.org/10.1016/j.clinph.2014.11.021

Ma, G., Troxel, A. B., & Heitjan, D. F. (2005). An index of local sensitivity to nonignorable drop-out in longitudinal modelling. Statistics in Medicine, 24(14), 2129–2150. https://doi.org/10.1002/sim.2107

Micanovic, C., & Pal, S. (2014). The diagnostic utility of EEG in early-onset dementia: A systematic review of the literature with narrative analysis. Journal of Neural Transmission, 121(1), 59–69. https://doi.org/10.1007/s00702-013-1070-5

Pijnenburg, Y. A. L., Strijers, R. L. M., Made, Y.vd, van der Flier, W. M., Scheltens, P., & Stam, C. J. (2008). Investigation of resting-state EEG functional connectivity in frontotemporal lobar degeneration. Clinical Neurophysiology, 119(8), 1732–1738. https://doi.org/10.1016/j.clinph.2008.02.024

Pinheiro J, Bates D, DebRoy S, Sarkar D, R Core Team (2020). nlme: Linear and Nonlinear Mixed Effects Models. R package version 3.1-148, <URL: https://CRAN.R-project.org/package=nlme>.

Pollitt, A., & Hutchinson, C. (1987). Calibrating graded assessments: Rasch partial credit analysis of performance in writing. Language Testing, 4(1), 72–92. https://doi.org/10.1177/026553228700400107

Ramsey, F. C. (1979). Protein-Energy Malnutrition in Barbados: The Role of Continuity of Care. New York, NY: Josiah Macy Jr. Foundation,121–122.

Roks, G., Korf, E. S. C., Van Der Flier, W. M., Scheltens, P., & Stam, C. J. (2008). The use of EEG in the diagnosis of dementia with Lewy bodies. Journal of Neurology, Neurosurgery and Psychiatry, 79(4), 377– 380. https://doi.org/10.1136/jnnp.2007.125385

Sam, M. C., & So, E. L. (2001). Significance of epileptiform discharges in patients without epilepsy in the community. Epilepsia, 42(10), 1273–1278.

Segalowitz, S. J., Santesso, D. L., and Jetha, M. K. (2010). Electrophysiological changes during adolescence: a review. Brain Cogn. 72, 86–100. doi: 10.1016/jbandc.2009.10.00

Shelley, B. P., Trimble, M. R., & Boutros, N. N. (2008). Electroencephalographic cerebral dysrhythmic abnormalities in the trinity of nonepileptic general population, neuropsychiatric, and neurobehavioral disorders. The Journal of neuropsychiatry and clinical neurosciences, 20(1), 7–22.

Taboada-Crispi, A., Bringas-Vega, M.L., Bosch-Bayard, J., Galán-García, L., Bryce, C., Rabinowitz, A.G., Prichep, L.S., Isenhart, R., Calzada-Reyes, A., Virues-Alba, T. and Guo, Y., 2018. Quantitative EEG tomography of early childhood malnutrition. Frontiers in neuroscience, 12, p.595.

Uhlhaas, P. J., Roux, F., Rodriguez, E., Rotarska-Jagiela, A., and Singer, W. (2010). Neural synchrony and the development of cortical networks. Trends Cogn. Sci.14, 72–80. doi: 10.1016/j.tics.2009.12.00

United Nations Development Program (UNDP)., 2016. UNDP Support to the Implementation of the 2030 Agenda for Sustainable Development.

Waldemar, G., Dubois, B., Emre, M., Georges, J., McKeith, I.G., Rossor, M., Scheltens, P., Tariska, P. and Winblad, B., 2007. Recommendations for the diagnosis and management of Alzheimer’s disease and other disorders associated with dementia: EFNS guideline. European Journal of Neurology, 14(1), pp.e1–e26.

Xie, H., Gao, W., Xing, B., Heitjan, D. F., Hedeker, D., & Yuan, C. (2018). Measuring the Impact of Nonignorable Missingness Using the R Package isni. Computer Methods and Programs in Biomedicine, 164, 207–220. https://doi.org/10.1016/j.cmpb.2018.06.014

